# Histopathological findings in COVID-19 cases: A Systematic Review

**DOI:** 10.1101/2020.10.11.20210849

**Authors:** Hamed Hammoud, Ahmed Bendari, Tasneem Bendari, Iheb Bougmiza

**Author notes:** **Corresponding author** Hamed Hammoud, MD, Community medicine department, Department of Medical Education, Hamad Medical Corporation.

## Abstract

**Background:** The current COVID-19 pandemic is considered one of the most serious public health crisis over the last few decades. Although the disease can result in diverse, multiorgan pathology, there have been very few studies addressing the postmortem pathological findings of the cases. Active autopsy amid this pandemic could be an essential tool for diagnosis, surveillance, and research.

**Objective:** To provide a total picture of the SARS-CoV-2 histopathological features of different body organs through a systematic search of the published literature.

**Methods:** A systematic search of electronic databases (PubMed, ScienceDirect, Google scholar, Medrxiv & Biorxiv) was carried out from December 2019 to August, 15^th^ 2020, for journal articles of different study designs reporting postmortem pathological findings in COVID-19 cases. PRISMA guidelines were used for reporting the review.

**Results:** A total of 50 articles reporting 430 cases were included in our analysis. Postmortem pathological findings were reported for different body organs, pulmonary system (42 articles), cardiovascular system ( 23 articles), hepatobiliary system (22 articles), kidney (16 articles), spleen, and lymph nodes (12 articles), and central nervous system (7 articles). In lung samples, diffuse alveolar damage (DAD) was the most commonly reported findings in 239 cases (84.4%). Myocardial hypertrophy (87 cases by 51.2%), arteriosclerosis (121 cases by 62%), and steatosis ( 118 cases by 59.3%) were the most commonly reported pathological findings in the heart, kidney, and hepatobiliary system respectively.

**Conclusion:** Autopsy examination as an investigation tool could help in a better understanding of SARS-CoV-2 pathophysiology, diagnosis, management, and subsequently improving patient care.

## 1. Introduction

The novel coronavirus disease (COVID-19) pandemic is considered one of the most challenging public health crisis in the past century. It first emerged in Wuhan, China, in late December 2019 and believed to be caused by infection with the severe acute respiratory syndrome coronavirus 2 (SARS- CoV- 2) virus. ^(1)^ The first cases of COVID-19 in China were believed to be of zoonotic origin, but the global spread of the disease was mainly travel-related. The disease has spread from China to affect nearly 200 countries all over the world. ^(2)^ The virus is easily transmissible via droplets and fomites or when bodily fluids of the infected individual come in contact with another person’s face, mouth, eyes, or nose. ^(3)^

Regarding pathogenesis, Angiotensin-converting enzyme 2 (ACE2), which is highly expressed on the respiratory tract, acts as a receptor to SARS-CoV-2. The virus invades the human cells causing massive destruction and inflammation of different organs and subsequently affecting the vascular supply and even progression to fibrosis. ^(4)^ The main clinical manifestations include fever, cough, fatigue, and shortness of breath. Other less common symptoms include headache, sore throat, and rhinorrhea. Along with that, one-fifth of patients (20%) presented with severe symptoms such as respiratory failure, multiorgan failure, septic shock, all of which necessitate intensive care. ^(5)^ Although COVID-19 is mainly affecting the respiratory system, there have been reported cases of cardiogenic and renal involvement in patients without previously known heart or renal diseases. ^(6, 7)^

The case-fatality rate for COVID-19 is variable across different nations, between 11.75% in Italy and 0.37% in South Africa. The mean recovery time is two weeks for mild cases and 3-6 weeks for severe or critical cases. ^(8)^ The diagnosis of COVID-19 relies mainly on reverse-transcription polymerase chain reaction (RT-PCR) with some emerging evidence on characteristic CT and laboratory findings. ^(9)^ COVID-19 is a member of the coronavirus family, including MERS-CoV and SARS-CoV. ^(10)^ Both MERS-CoV and SARS-CoV are believed to affect humans and cause interstitial pneumonia, pneumocyte hyperplasia, and acute diffuse alveolar damage. ^(11, 12)^ The diverse histopathological findings associated with COVID-19 infections suggest that multiple organs are affected by the virus, with the pulmonary system is the most common system to be affected. Carsana et al. showed in their study the variety of the pathological findings of COVID-19 in the respiratory system. They’ve found that the pneumocytes desquamation, pulmonary edema, and diffuse alveolar damage are the most common microscopic findings. ^(13)^

As of August 10^th,^ 2020, the number of COVID-19 cases worldwide surpassed 20,162,474 million cases, with almost 737,417 deaths. However, the number of studies addressing the postmortem autopsy findings of COVID-19 patients is still scarce compared to the number of deaths. This could be explained by the fears of contagiousness associated with COVID-19 infection.

Since the beginning of the pandemic, the Centers for Disease Control and Prevention (CDC) released interim guidelines for the collection and analysis of clinical specimens that might contain SARS-CoV-2. ^(14)^

Active autopsy amid emerging epidemic diseases has been identified as an essential tool for diagnosis, surveillance, and research. Pathologists are usually among the first health care professionals in identifying novel infectious agents outbreaks. ^(15)^ Our aim in this systematic review is to provide a total picture of the SARS-CoV-2 histopathological features of different body organs in postmortem autopsies through a systematic search of the published literature. We believe this will help in a better understanding of mechanisms of injury and pathophysiology of severe SARS-CoV-2 infection and subsequently improving patient care.

## 2. Methods

This study followed the recommendations established by the Preferred Reporting Items for Systematic Reviews and Meta-Analyses (PRISMA) statement. (Appendix-1) ^(16)^

### 2.1. Sources of information

A predetermined protocol was used to perform this systematic review using the following databases: PubMed, Google Scholar, ScienceDirect, and MedRxiv. The search included articles published between December 2019 and August 15th, 2020. The reference lists of relevant studies were hand-searched to identify cited articles that were not captured by electronic search.

### 2.2. Selection criteria

Articles were included if they met the following eligibility criteria: (1) addressed pathological reports of COVID-19 autopsies or postmortem cases, (2) involved human subjects (at least one case), (3) all study designs were involved (case report, case series, cross-sectional, case-control, randomized and non-randomized studies), (4) no language restrictions were applied.

### 2.3. Study selection and search terms

The search terms and keywords across the different databases have been provided in (Appendix-2). The selection was broad to include as many studies as possible. In the initial phase, two independent reviewers (H.H. & A.B.) screened the titles and abstracts of the articles using the Rayyan QCRI^®^ website. As a result, all non-relevant articles were excluded. In the second phase, the full-texts of the remaining articles were independently reviewed for the final selection of eligible studies. Any disagreement between the two reviewers was resolved by a third reviewer (T.B.).

### 2.4. Quality assessment and risk of bias

To assess the internal validity of the included studies, we used different tools according to study design. For cross-sectional studies, the Newcastle-Ottawa Quality Assessment Scale (NOS) (modified for cross-sectional studies) was used after removing items that relate to comparability and adjustment. The tool contains three major subsections (Selection, Comparability, and Outcome). A score for quality, modified from the tool, was used to assess the appropriateness of study design, recruitment strategy, sample representativeness, reliability of the outcome, sample size provided, and appropriate statistical analyses. At least two reviewers (H.H., A.B., T.B.) independently ranked these domains. When the independent evaluations of the ranks differed between the two reviewers, they discussed disagreements to reach for mutual decision. For case reports and case series, a version of (NOS) checklist was adapted by Murad et al. to assess the methodological quality of case reports and case series. ^(17)^ By this approach, we assessed the quality of each study with regard to four domains: selection, ascertainment; causality; and reporting. From the results of each checklist, if 25% or less of the criteria were addressed, the article was scored as *poor*; if 26% to 50% of the criteria were addressed, the article was scored as *fair*; if 51% to 75% of criteria were addressed, the article was scored as *good*; and if 76% to 100% of the criteria were addressed, the article was scored as *excellent*.

### 2.5. Data extraction

A single author (A.B.) extracted the variables from each included study. The data from the final list of included articles onto an online *Google sheet*. Several study characteristics were extracted, including;

- General characteristics as study type, country of origin, article language sample size.
- Study population demographics like age and gender.
- Clinical findings like symptomatology, lab findings, and patient comorbidities.
- Histopathologic and microscopic findings of different organs.

## 2. Results

*Figure 1* is a flow chart showing the procedure for the selection of studies. Initially, we identified 3297 from 5 databases as follows; PubMed (2,262), ScienceDirect (189), MedRxiv & BioRxiv (71), and Google Scholar (775). After the initial title screening, there were 689 articles by **08/15/2020**. 211 duplicates were removed, leaving 478 for abstract screening. After excluding non-relevant articles, the final number of included articles is (50).

**Figure (1):**
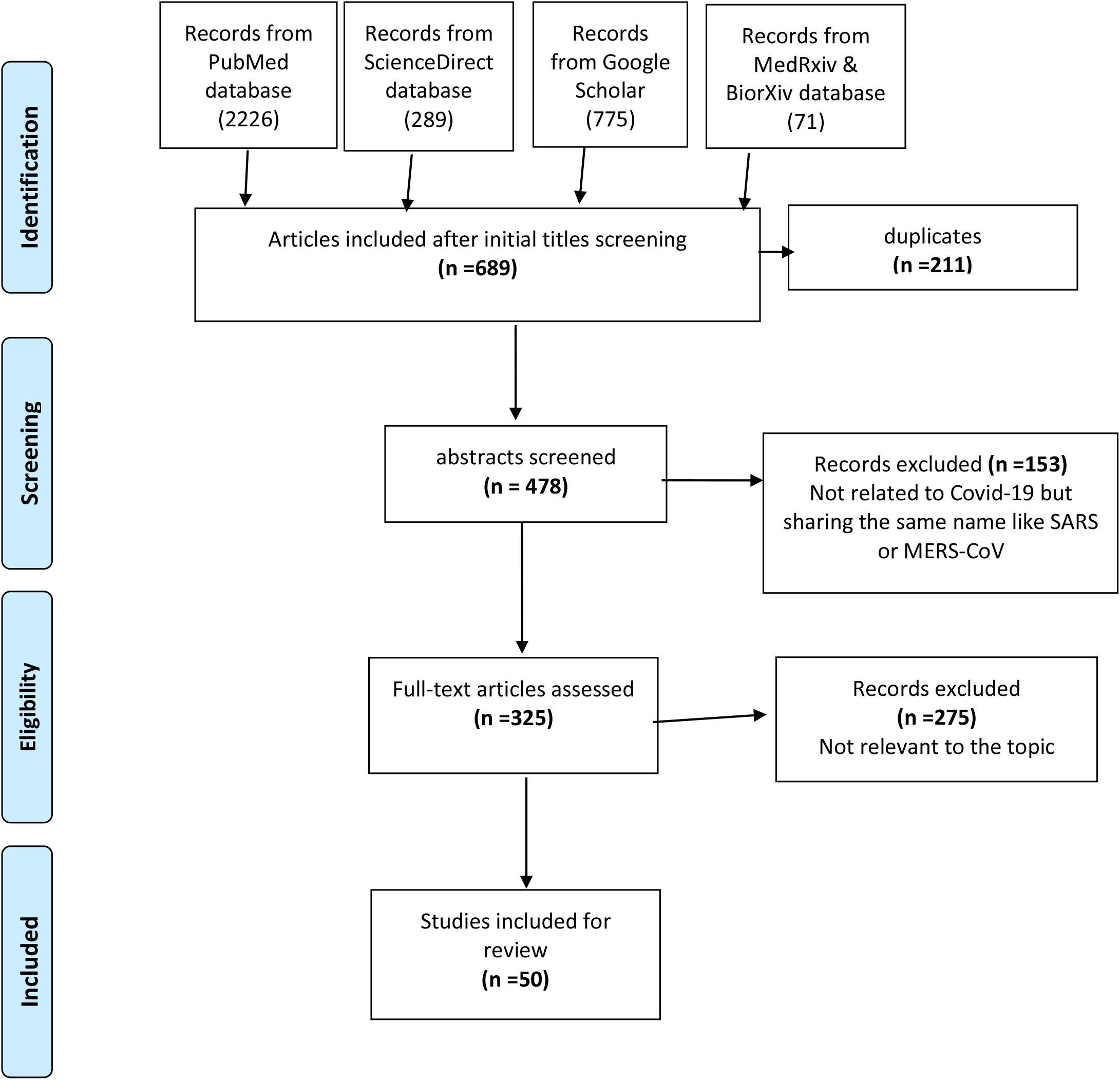
Flow chart showing the procedure for selection of studies.

### 3.1 Characteristics of included studies

A total number of 50 studies were included in our systematic review, with overall 430 cases. Regarding the date of publication, only three studies were published in February, two in March, seven in April, and started to increase to reach 15 in May, which is the month with the highest number of publications. *(Figure 2)* Regarding the type of published articles, case reports and case series were the most common type by 39 studies, then cross-sectional studies with 10 publications, and 1 cohort study. Regarding the country of origin of the published studies, there were about 16 countries. The USA was the country with the most publications (16 studies), followed by China (10 studies), Germany (6 studies), Italy (5 studies), Switzerland (2 studies), and one study for each of the following countries (Iran, Finland, Austria, Belgium, Japan, Spain, Netherland, UK, Romania, Austria, & Denmark). Regarding the Language of the published articles, there were only two languages English & Chinese. English with 47 studies and Chinese with 3 studies. *(Table 1)*

**Figure (2):**
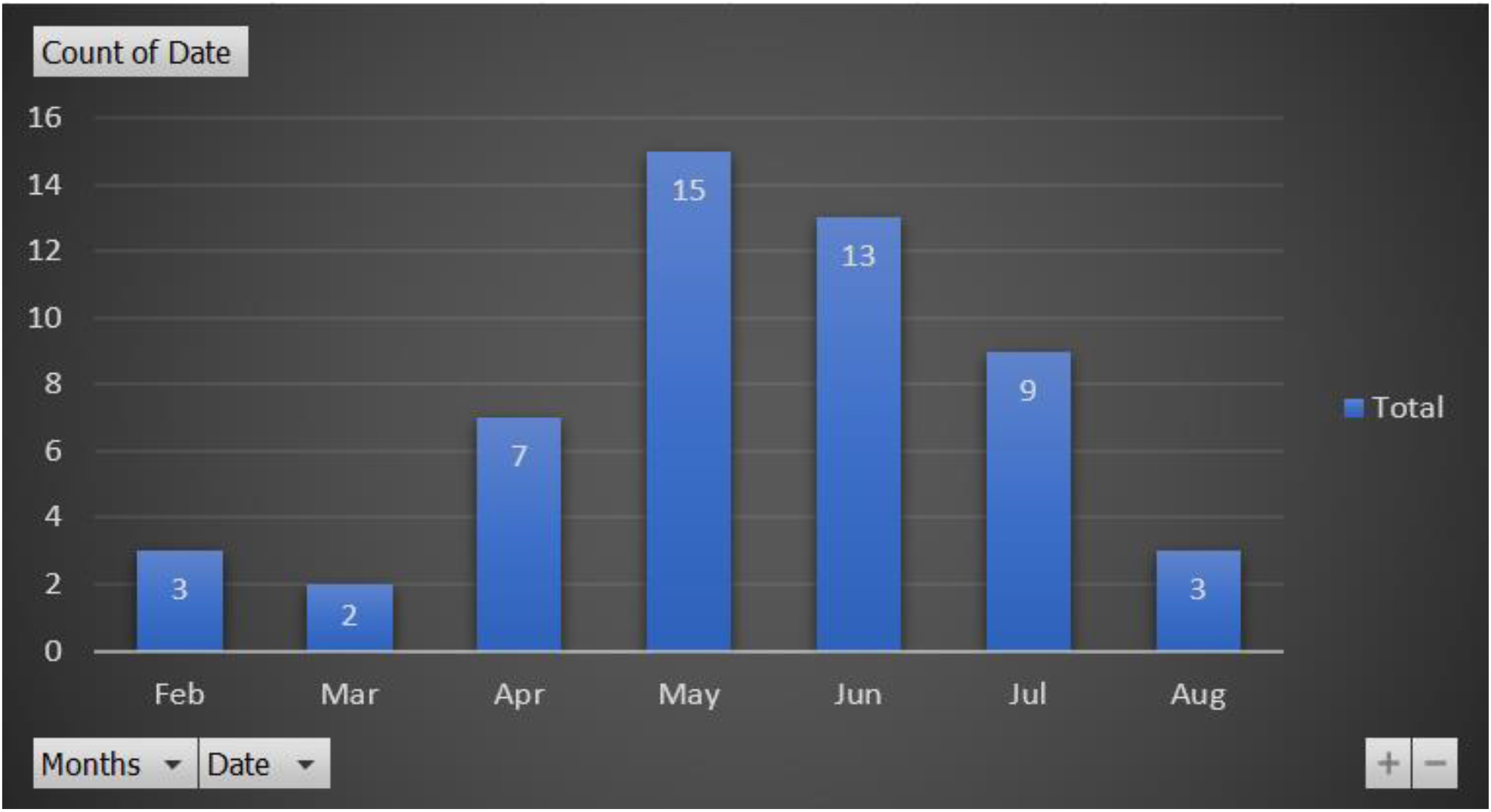
Timeline distribution of published articles.

### 3.2 Quality of evidence

We used the GRADE framework for judging the precision and confidence estimate in the review. Generally speaking, the evidence derived from observational studies is classified as of low quality. ^(18)^ Regarding the risk of bias assessment in the review, 4 articles scored between 26 -50 %, which is considered “Fair” ^(19-22)^ 26 articles scored between 51-75 %, which is considered as “Good” ^(13, 23-47)^, and 20 articles scored more than 76 % which is considered as “Excellent.” ^(48-67)^ A high degree of inconsistency was noticed in the review as the study populations were somehow heterogeneous in the main characteristics like age, gender, and comorbidities. Although there were no language restrictions applied in the review, publication bias may appear due to the fact that the number of the published literature was small, especially at the beginning of the pandemic. Moreover, a very small number of countries were reporting autopsy findings. Regarding the indirectness, the majority of included studies used the same tool in diagnosing COVID-19, which is (RT-PCR), the same tool in identifying histopathological findings, and the studied population varied between studies. Hence, the quality of evidence was rated as “Moderate.” (Appendix-4)

### 3.3 Clinical findings of the cases

The review described a total of 430 patients with COVID-19. Among the included patients, gender was reported in 349 patients as follows; 297 males (85.1%) and 133 (14.9 %) female. Among the no patients for whom age was reported, the median age was (range: 11 to 94 years). Regarding the presenting symptoms of patients whom clinical symptoms were reported (192 patients), fever was reported in 121 patients followed cough in 103 patients and dyspnea (91 patients). ^(19-26, 28-32, 34, 36, 38-42, 44-48, 51-53, 55-57, 59, 60, 62-66)^ Regarding the pre-existing co-morbidities in patients whom medical history was reported, hypertension was found in 210 patients (48.8%), followed by coronary heart disease in 190 (44%), Diabetes in 134 patient (31%), chronic kidney disease in 96 patients (22.3%), obesity in 64 patients (14.8%), chronic lung disease in 54 patients (12.5%) and cancer in 50 patients (11.6%). ^(13, 19, 20, 22-32, 34, 38-41, 44-65, 67)^ Regarding the included organs, this review described the histopathology of different organs as follows; Lung and pulmonary system was the most common described organ in 42 articles, ^(13, 19-26, 28-32, 34-37, 39-41, 44-46, 48-60, 62-66)^ followed by heart in 23 articles ^(19, 20, 23, 24, 26, 28, 29, 31, 32, 40, 41, 44-46, 48-52, 54, 63, 65, 66)^, Liver in 21 articles ^(19, 20, 23, 24, 26, 28, 29, 31, 41, 44, 45, 48-52, 54, 61-63, 65)^, Kidney in 16 articles, ^(20, 23, 27-29, 31, 46, 48-52, 54, 56, 65, 67)^ spleen and lymph nodes in 12 articles, ^(20, 28, 29, 31, 33, 38, 49, 50, 52, 54, 56)^ CNS in 7 articles, ^(19, 29, 45, 47, 51, 54, 64)^ Skin in 2 articles ^(43, 53)^ gall bladder (1 article), ^(42)^ Pharynx (1 article). ^(50)^

### 3.4 Laboratory investigations

In all included studies, RT-PCR of Nasopharyngeal swab was the main method to confirm the positivity of COVID-19 in all patients. RT-PCR for COVID-19 of the Endotracheal Aspirate was reported in one patient. ^(66)^ RT-PCR for COVID-19 of the skin biopsy was reported in 7 patients. ^(43)^ Chest imaging, whether CT or CXR, was also reported in 206 patients (47.9%). ^(13, 19, 21, 23-32, 34, 39, 44, 46, 47, 50, 52, 53, 55-59, 62, 63, 66)^ Post-mortem RT-PCR for COVID-19 of the lung parenchyma was reported in 91 cases (21%). ^(22, 23, 29, 31, 34, 36, 39, 40, 45, 46, 50, 51, 57, 63, 65)^

### 3.5 Lung histopathological findings

In the 42 articles that described lung pathology. The most common reported pathological findings were diffuse alveolar injury in 239 cases (84.4%), ^(13, 19-26, 28-32, 34-37, 39-41, 44-46, 48-51, 54-60, 62-66)^ followed by hyaline membrane formation in 184 cases (65%), ^(13, 19-24, 26, 28, 30, 32, 35, 36, 39-41, 44-46, 48-57, 60, 62, 63, 66)^ Lymphocyte and/or monocyte infiltrates in 172 cases (60.7%), ^(13, 19, 20, 23-26, 28-30, 32, 34-36, 39-41, 44-46, 48-55, 57, 60, 63, 65, 66)^ pneumocyte hyperplasia in 171 cases (60.4%), ^(13, 19, 21, 25, 26, 28, 29, 34, 36, 39-41, 44-46, 48, 50-60, 63, 65)^ pulmonary microthrombi in 151 cases (53.3%), ^(13, 20-22, 28, 30-32, 35-37, 45, 48, 50-52, 54, 55, 57-60, 64, 65)^ fibrin exudation in 112 cases (39.8%), ^(13, 20, 21, 28, 32, 34, 36, 39-41, 44, 46, 48, 50, 51, 53, 55, 57, 60, 62)^ lung fibrosis in 97 cases (34.2%), ^(13, 19, 25, 29, 30, 32, 34, 37, 48, 50, 51, 55-57, 62, 66)^ intra-alveolar neutrophilic infiltration in 92 cases (32.5%), ^(13, 19, 21, 23, 26, 28, 30, 32, 36, 39, 41, 46, 48-50, 53, 57, 60)^ intra-alveolar hemorrhage in 86 cases (30.4%), ^(13, 26, 28, 29, 31, 32, 41, 44, 46, 48, 49, 51, 53, 56, 65)^ interstitial thickening in 80 cases (28.3%), ^(13, 19, 25, 26, 29, 31, 34, 36, 40, 48, 55, 58, 60, 65)^ vascular congestion in 77 cases (27.2%), ^(13, 20, 23, 25, 26, 28, 30, 39, 41, 45, 48, 49, 53, 56)^ pneumocyte damage in 73 cases (25.8%), ^(13, 20, 24, 26, 32, 34, 46, 48, 58, 60, 62)^ squamous metaplasia in 68 cases (24%), ^(13, 19, 28, 30, 31, 37, 44, 46, 50, 56, 59, 60, 66)^ viral inclusion in 45 cases (15.9%), ^(13, 25, 29, 31, 48, 54, 60)^ serous exudation in 19 cases (6.7%), ^(46, 48, 51, 58)^ fibrinoid vascular necrosis in 17 cases (6%), ^(19, 26, 44, 49, 53, 60)^ and pulmonary embolism in 14 cases (4.9%). ^(28, 29, 40, 49, 54, 57)^ (Appendix-3)

### 3.6 Heart histopathological findings

In the 23 articles that described heart pathology. The most reported pathology was myocardial hypertrophy in 87 cases (51.2%), ^(26, 28, 29, 31, 48, 49, 51, 52, 54)^ followed by myocardial fibrosis in 85 cases (50%), ^(26, 28, 29, 31, 32, 40, 45, 46, 50-52, 54)^ coronary small vessel disease in 44 cases (25.9%) ^(20, 23, 28, 40, 49, 52, 54, 63)^ myocardial cell infiltrate in 27 cases (15.9%), ^(19, 20, 24, 44, 48, 50, 52, 54, 65)^ cardiac amyloidosis in 10 cases (5.9%), ^(29, 49, 52)^ and myocardial necrosis in 9 cases (5.3%). ^(44, 46, 48, 49)^ (Appendix-3)

### 3.7 Liver histopathological findings

In the 21 articles that described liver pathology. The most reported pathology was steatosis in 118 cases (59.3%), ^(20, 23, 24, 26, 28, 29, 31, 33, 41, 44, 45, 49, 51, 52, 54, 61-63, 65)^ followed by fibrosis in 62 cases (31.1%), ^(19, 28, 52, 54, 61, 65)^ hepatic congestion in 59 cases (29.6%), ^(29, 33, 44, 50-52, 65)^ cellular infiltrate in 54 cases (27.1%), ^(19, 24, 26, 29, 52, 61, 62, 65)^ hepatic necrosis in 44 cases (22.1%), ^(20, 26, 29, 41, 44, 48, 52, 61, 62)^ cholestasis in 8 cases (4%), ^(52)^ and cirrhosis in 4 cases (2%). ^(26, 51)^ (Appendix-3)

### 3.8 Kidney histopathological findings

In the 16 articles that described kidney pathology. The most reported pathology was arteriosclerosis in 121 cases (62%), ^(23, 27-29, 50, 54, 67)^ followed by nephrosclerosis in 91 cases (46.7%), ^(23, 29, 54, 67)^ acute tubular injury in 86 cases (44.1%), ^(20, 27, 31, 48, 49, 52, 54, 65, 67)^ glomerulosclerosis in 70 cases (35.9%), ^(27-29, 31, 46, 51, 52)^ tubular cast in 38 cases (19.5%), ^(20, 27, 29, 48, 51)^ glomerular fibrin thrombus in 21 cases (10.7%), ^(20, 27-29, 54, 56, 65, 67)^, and viral particles in 16 cases (8.2%). ^(20, 27, 28)^ (Appendix-3)

### 3.9 Immune system (Spleen and lymph node) histopathological findings

In the 12 articles that described spleen pathology. The most reported pathology was lymphocyte depletion in 38 cases (31.4%), ^(20, 28, 29, 38, 48, 52, 54)^ followed by hemophagocytosis in Spleen in 12 cases (9.9%). ^(33, 38, 54, 56)^ In the 11 articles that described lymph node pathology, the most common pathology that had been reported was lymphocyte depletion in 23 cases (20.7%), ^(20, 33, 48, 52, 54)^ hemophagocytosis in the lymph nodes in 22 cases (19.8%). ^(29, 31, 33, 54, 56)^

### 3.10 CNS histopathological findings

In the 7 articles that described CNS pathology. The most common reported pathology was cerebral hemorrhage in 11 cases (15.5%), ^(29, 47, 51, 64)^ focal spongiosis in 11 cases (15.5%), ^(47, 51)^ and vascular congestion in 11 cases (15.5%), ^(51, 54)^ followed by diffuse or focal ischemic necrosis in 9 cases (12.7%). (51, 54)

### 3.11 Skin histopathological findings

In the 2 articles that described skin pathology, the most common reported pathology was thrombogenic vasculopathy in 4 cases (10,0.4%), ^(43, 53)^ followed perivascular inflammation in 2 cases (10,0.2%) ^(43, 53)^ and vasculitis in one case ^(43)^

### 3.12 Gall bladder histopathologic findings

In the one article that described gall bladder postmortem pathology, inflammatory infiltration and endoluminal obliteration of vessels with wall breakthrough, hemorrhagic infarction, and nerve hypertrophy were reported in one case report. ^(42)^

### 3.13 Pharynx histopathological findings

One study described pharyngeal postmortem pathology. The study included 8 cases, 7 of which reported mild to pronounced lymphocytic pharyngitis. ^(50)^

## 4. Discussion

This systematic review identified 50 studies with a total of 430 patients and postmortem pathological findings of different body organs. Since the beginning of the pandemic, the body of evidence and the number of published studies have increased over time, but it is still limited compared to the number of COVID-19 deaths almost (1 million deaths). There were only 16 countries that contributed in publishing autopsy reports of the COVID-19 deaths, which is considered very low considering the fact that the disease affected nearly 200 countries all over the world. ^(2)^ With regard to the timeline of the published studies, it took us five months (up to May 2020) so that we can find an appropriate number of publications addressing the postmortem pathological findings. One of the main reasons for the limited published literature is the fear of COVID-19 infection transmission during the postmortem examination and the perceived risk of healthcare professionals, especially pathologists, about this ‘new’ disease with a poor understanding of its pathological mechanism, especially at the beginning of the pandemic. ^(68)^ Moreover, in some countries, the number of safe autopsy rooms is very low, which, according to the WHO & CDC guidelines, is considered one of the barriers that contributed to the scarce evidence. ^(69-71)^

### Postmortem pulmonary findings

Regarding the postmortem pulmonary pathology, our review showed that different histopathological findings had been identified among COVID-19 cases. Diffuse alveolar injury, hyaline membrane formation, pneumocyte hyperplasia, microthrombi, fibrin exudation, pulmonary fibrosis, and intra-alveolar hemorrhage are among the most frequently reported pathological findings. Diffuse alveolar damage (DAD) has been reported the most among all pulmonary findings with 239 cases (84 %). On second thoughts, these pathological findings should not be seen as one of the COVID-19 attributes without considering other important factors affecting the course of illness like age, symptoms, comorbidities, and management plan. For example, Wichmann et al. investigated the possibility of attributed venous thromboembolism in a cohort of COVID-19 patients. Autopsy findings found that deep venous thrombosis was found in 58 % of the cases, and this has been linked to COVID-19 while ignoring the fact that the majority of patients suffered from atrial fibrillation, coronary heart disease, and cancers, which have been proven to be decisive determinant factors in developing thromboembolism. ^(30)^

On the other hand, similar pathological and autopsy findings have been reported in deceased patients with other coronavirus infections such as Middle East respiratory syndrome coronavirus (MERS-CoV) and severe acute respiratory syndrome (SARS). NG et al. published a case report of a 45 years old patient with MERS-CoV with autopsy findings after death. Postmortem pulmonary findings included diffuse alveolar damage, type 2 pneumocyte hyperplasia, interstitial infiltrate, alveolar fibrin deposits, and prominent hyaline membranes. ^(72)^ Alsaad et al. also reported DAD in his report that included 33 years old patient with MERS-CoV. ^(11)^ Moreover, Franks et al. reported in their study of 8 SARS patients that DAD was the main pathological findings. In contrast, Nicholas et al. reported other findings like the focal deposition of fibrin along the exposed basement membrane. ^(12, 73)^ On the other hand and in non-coronaviruses pulmonary infection like H1N1 cases, histopathological findings as septal inflammation, congestion, and thickening of alveolar septae, patchy peripheral hemorrhage, and diffuse alveolar hemorrhage have been reported by different studies. ^(74-77)^

### Other Organ findings

Regarding the postmortem cardiac pathology, there were 23 studies with a total number of 87 cases addressing the histopathological findings in the heart. Myocardial hypertrophy, small coronary vessels, cardiac fibrosis, cardiac cell infiltrate, and cardiac amyloidosis are the main findings. Although viral myocarditis has been reported in patients with SARS-Cov-2 virus, lymphocyte infiltrate was found only in one case reported by Buja et al. during Immunohistochemical (IHC) staining. ^(28, 78)^ These pathological findings could be attributed to the comorbidities of affected patients, as most of them suffered from hypertension, diabetes, or coronary heart disease. On the other hand, myocardial edema and fibrosis have been recorded in deceased patients with SARS and MERS-CoV. ^(72, 79, 80)^ As the studies in this review reported that nephrosclerosis, arteriosclerosis, glomerulosclerosis, and acute tubular injury were the most commonly reported findings in the postmortem renal biopsies, other pathological findings like hyaline arteriolosclerosis, patchy interstitial inflammation, and granular casts have been reported in other coronavirus cases like SARS & MERS-CoV. ^(72, 81, 82)^ Regarding the pathological findings in the hepatobiliary system, our review found that hepatic fibrosis, steatosis, cirrhosis, and interstitial inflammations were the main findings. In contrast, other pathological findings were reported in patients with SARS-CoV-1 infection like lymphocytic infiltrate and balloon degeneration. ^(83)^ As for histopathological findings of the spleen and lymph nodes, lymphocyte depletion and hemophagocytosis of the Spleen and lymph nodes were the main findings. Our results were consistent with similar pathological findings from other coronavirus infections. ^(79, 84)^

Although the SARS-CoV-2 virus hasn’t been detected in the spinal fluid, our study suggests that COVID-19 had been capable of infecting the central nervous system via olfactory and trigeminal nerves causing cerebral hemorrhage, focal spongiosis, and vascular congestion. ^(85)^ On the contrary, and in the case of SARS infection, RT-PCR has detected the genomic sequences of the virus in cerebral spinal fluid and brain tissue specimens and was responsible for brain edema and neuronal degeneration. ^(81, 86)^

### Limitations of the study

As a part of any research, we faced many limitations while conduction the review. First, in this study, we focused on the available studies in certain databases in the first months of the pandemic, so government reports and other relevant grey literature weren’t included in this review, so publication bias is a possibility. Second, due to the scarcity of the evidence, we decided to include pre-prints. These publications have not yet undergone peer review. However, since we assessed the risk of bias of these studies, we feel that the benefits of including the data from these pre-prints in our review outweigh the risks. Third, we’ve included only 50 articles, but we can’t ignore the fact that the number of publications is increasing daily, and we might have missed the recently published ones. Fourth, missing information in some of the published articles has been a challenge. Many articles didn’t report the basic characteristics of the cases like gender, comorbidities, and clinical course of the disease.

## 5. Conclusion

Postmortem histopathological biopsies play an essential role in understanding the pathophysiology of SARS-CoV-2 infection. COVID-19 affects different body organs with different pathological features throughout the course of infection. Cellular destruction, vascular invasion, and fibrous formation have been identified in the pulmonary, hepatobiliary, and renal systems, while they are not clear in other systems like CNS and genitourinary system. Further research is needed for a better understanding of the disease and exploring the extent of its effects on different organs and tissues.

## Data Availability

Tables
https://www.editorialmanager.com/prp/download.aspx?id=124729&guid=0c9e8fd8-0abe-4b95-bd5a-13d27daaf816&scheme=1
appendix 1
https://www.editorialmanager.com/prp/download.aspx?id=124731&guid=f8ebb7b6-363d-4636-8d76-8e20816f49da&scheme=1
Appendix 2
https://www.editorialmanager.com/prp/download.aspx?id=124732&guid=7cb1ad8e-32fb-4aec-97b6-c8606d7583d0&scheme=1
Appendix 3
https://www.editorialmanager.com/prp/download.aspx?id=124733&guid=e2671aff-6beb-4d7e-81dc-6505e413768a&scheme=1
Appendix 4
https://www.editorialmanager.com/prp/download.aspx?id=124734&guid=e1be8ed9-9d68-4026-b781-6f4c071f11eb&scheme=1

## Ethical

No ethical approval is required.

## Patient Consent

Not required.

## References

1. Wu F, Zhao S, Yu B, Chen Y-M, Wang W, Song Z-G, et al. A new coronavirus associated with human respiratory disease in China. 2020;579(7798):265–9.

2. Organization WH. Rolling updates on coronavirus disease (COVID-19) [Internet] Geneva, Switzerland 2020 [updated Updated 31 July 2020Accessed 08-09-2020]. available from: https://www.who.int/emergencies/diseases/novel-coronavirus-2019/events-as-they-happen.

3. Jayaweera M, Perera H, Gunawardana B, Manatunge J. Transmission of COVID-19 virus by droplets and aerosols: A critical review on the unresolved dichotomy. (1096-0953 (Electronic)).

4. Lukassen SA-O, Chua RL, Trefzer T, Kahn NC, Schneider MA, Muley T, et al. SARS-CoV-2 receptor ACE2 and TMPRSS2 are primarily expressed in bronchial transient secretory cells. (1460-2075 (Electronic)).

5. Li LQ, Huang TA-O, Wang YQ, Wang ZP, Liang Y, Huang TB, et al. COVID-19 patients’ clinical characteristics, discharge rate, and fatality rate of meta-analysis. (1096-9071 (Electronic)).

6. Zhang H, Liao YS, Gong J, Liu J, Xia X, Zhang H. Clinical characteristics of coronavirus disease (COVID-19) patients with gastrointestinal symptoms: A report of 164 cases. LID - S1590-8658(20)30189-4 [pii] LID -10.1016/j.dld.2020.04.034 [doi]. (1878-3562 (Electronic)).

7. Khan IH, Zahra SA, Zaim S, Harky Ajjocs. At the heart of COVID-19. 2020.

8. Giorgi Rossi P, Broccoli S, Angelini P. Case fatality rate in patients with COVID-19 infection and its relationship with length of follow up. (1873-5967 (Electronic)).

9. Rodriguez-Morales AJ, Cardona-Ospina JA, Gutiérrez-Ocampo E, Villamizar-Peña R, Holguin-Rivera Y, Escalera-Antezana JP, et al. Clinical, laboratory and imaging features of COVID-19: A systematic review and meta-analysis. (1873-0442 (Electronic)).

10. Liu M, Feng RE, Li Q, Zhang HK, Wang YG. [Comparison of pathological changes and pathogenic mechanisms caused by H1N1 influenza virus, highly pathogenic H5N1 avian influenza virus, SARS-CoV, MERS-CoV and 2019-nCoV]. (0529-5807 (Print)).

11. Alsaad KA-O, Hajeer AH, Al Balwi M, Al Moaiqel M, Al Oudah N, Al Ajlan A, et al. Histopathology of Middle East respiratory syndrome coronovirus (MERS-CoV) infection clinicopathological and ultrastructural study. (1365-2559 (Electronic)).

12. Franks TJ, Chong Py Fau - Chui P, Chui P Fau - Galvin JR, Galvin Jr Fau - Lourens RM, Lourens Rm Fau - Reid AH, Reid Ah Fau - Selbs E, et al. Lung pathology of severe acute respiratory syndrome (SARS): a study of 8 autopsy cases from Singapore. (0046-8177 (Print)).

13. Carsana L, Sonzogni A, Nasr A, Rossi RS, Pellegrinelli A, Zerbi P, et al. Pulmonary postmortem findings in a series of COVID-19 cases from northern Italy: a two-centre descriptive study. The Lancet Infectious Diseases.

14. Centers for Disease Control and Prevention. Collection and Submission of Postmortem Specimens from Deceased Persons with Known or Suspected COVID-19 [Internet] USA 2020 [updated Updated June 4, 202013 August 2020]. available from: https://www.cdc.gov/coronavirus/2019-ncov/hcp/guidance-postmortem-specimens.html.

15. Schwartz DA, Herman CJ. Editorial Response: The Importance of the Autopsy in Emerging and Reemerging Infectious Diseases. Clinical Infectious Diseases. 1996;23(2):248–54.

16. Moher D, Liberati A, Tetzlaff J, Altman DG. Preferred reporting items for systematic reviews and meta-analyses: the PRISMA statement. BMJ. 2009;339:b2535.

17. Murad MH, Sultan S, Haffar S, Bazerbachi F. Methodological quality and synthesis of case series and case reports. BMJ Evidence-Based Medicine. 2018;23(2):60.

18. Schünemann H, Brozek, J., Guyatt, G., & Oxman, A. (Eds.). GRADE handbook for grading quality of evidence and strength of recommendations. Canada: The GRADE Working Group; 2013 [12-09-2019 [Updated October 2013]]. available from: https://gdt.gradepro.org/app/handbook/handbook.html.

19. Schaller T, Hirschbühl K, Burkhardt K, Braun G, Trepel M, Märkl B, et al. Postmortem Examination of Patients With COVID-19. JAMA. 2020;323(24):2518–20.

20. Rapkiewicz AV, Mai X, Carsons SE, Pittaluga S, Kleiner DE, Berger JS, et al. Megakaryocytes and platelet-fibrin thrombi characterize multiorgan thrombosis at autopsy in COVID-19: A case series. (2589-5370 (Electronic)).

21. Konopka KE, Wilson A, Myers JL. Postmortem Lung Findings in a Patient With Asthma and Coronavirus Disease 2019. CHEST. 2020;158(3):e99–e101.

22. Konopka KE, Nguyen T, Jentzen JM, Rayes O, Schmidt CJ, Wilson AM, et al. Diffuse Alveolar Damage (DAD) from Coronavirus Disease 2019 Infection is Morphologically Indistinguishable from Other Causes of DAD. LID - 10.1111/his.14180 [doi]. (1365-2559 (Electronic)).

23. Barton LM, Duval EJ, Stroberg E, Ghosh S, Mukhopadhyay S. COVID-19 Autopsies, Oklahoma, USA. American Journal of Clinical Pathology. 2020;153(6):725–33.

24. Xu Z, Shi L, Wang Y, Zhang J, Huang L, Zhang C, et al. Pathological findings of COVID-19 associated with acute respiratory distress syndrome. (2213-2619 (Electronic)).

25. Tian S, Hu W, Niu L, Liu H, Xu H, Xiao SY. Pulmonary Pathology of Early-Phase 2019 Novel Coronavirus (COVID-19) Pneumonia in Two Patients With Lung Cancer. (1556-1380 (Electronic)).

26. Tian S, Xiong Y, Liu H, Niu L, Guo J, Liao M, et al. Pathological study of the 2019 novel coronavirus disease (COVID-19) through postmortem core biopsies. (1530-0285 (Electronic)).

27. Su H, Yang M, Wan C, Yi LX, Tang F, Zhu HY, et al. Renal histopathological analysis of 26 postmortem findings of patients with COVID-19 in China. (1523-1755 (Electronic)).

28. Buja LM, Wolf DA, Zhao B, Akkanti B, McDonald M, Lelenwa L, et al. The emerging spectrum of cardiopulmonary pathology of the coronavirus disease 2019 (COVID-19): Report of 3 autopsies from Houston, Texas, and review of autopsy findings from other United States cities. (1879-1336 (Electronic)).

29. Bradley BT, Maioli H, Johnston R, Chaudhry I, Fink SL, Xu H, et al. Histopathology and ultrastructural findings of fatal COVID-19 infections in Washington State: a case series. (1474-547X (Electronic)).

30. Wichmann D, Sperhake JP, Lütgehetmann M, Steurer S, Edler C, Heinemann A, et al. Autopsy Findings and Venous Thromboembolism in Patients With COVID-19: A Prospective Cohort Study. (1539-3704 (Electronic)).

31. Martines Rb Fau - Ritter JM, Ritter Jm Fau - Matkovic E, Matkovic E Fau - Gary J, Gary J Fau - Bollweg BC, Bollweg Bc Fau - Bullock H, Bullock H Fau - Goldsmith CS, et al. Pathology and Pathogenesis of SARS-CoV-2 Associated with Fatal Coronavirus Disease, United States. (1080-6059 (Electronic)).

32. Fox SE, Akmatbekov A, Harbert JL, Li G, Quincy Brown J, Vander Heide RS. Pulmonary and cardiac pathology in African American patients with COVID-19: an autopsy series from New Orleans. (2213-2619 (Electronic)).

33. Prilutskiy A, Kritselis M, Shevtsov A, Yambayev I, Vadlamudi C, Zhao Q, et al. SARS-CoV-2 Infection Associated Hemophagocytic Lymphohistiocytosis: An autopsy series with clinical and laboratory correlation. medRxiv. 2020:2020.05.07.20094888.

34. Zhang H, Zhou P, Wei Y, Yue H, Wang Y, Hu M, et al. Histopathologic Changes and SARS-CoV-2 Immunostaining in the Lung of a Patient With COVID-19. (1539-3704 (Electronic)).

35. Li G, Fox SE, Summa B, Wenk C, Akmatbekov A, Harbert JL, et al. Multiscale 3-dimensional pathology findings of COVID-19 diseased lung using high-resolution cleared tissue microscopy. bioRxiv. 2020:2020.04.11.037473.

36. Cipolloni LA-O, Sessa FA-O, Bertozzi GA-O, Baldari B, Cantatore S, Testi R, et al. Preliminary Post-Mortem COVID-19 Evidence of Endothelial Injury and Factor VIII Hyperexpression. LID - E575 [pii] LID - 10.3390/diagnostics10080575 [doi]. (2075-4418 (Print)).

37. Grillo F, Barisione E, Ball L, Mastracci L, Fiocca R. Lung fibrosis: an undervalued finding in COVID-19 pathological series. LID - S1473-3099(20)30582-X [pii] LID - 10.1016/S1473-3099(20)30582-X [doi] FAU - Grillo, Federica. (1474-4457 (Electronic)).

38. Xu X, Chang XN, Pan HX, Su H, Huang B, Yang M, et al. [Pathological changes of the spleen in ten patients with coronavirus disease 2019(COVID-19) by postmortem needle autopsy]. (0529-5807 (Print)).

39. Wu JH, Li X, Huang B, Su H, Li Y, Luo DJ, et al. [Pathological changes of fatal coronavirus disease 2019 (COVID-19) in the lungs: report of 10 cases by postmortem needle autopsy]. (0529-5807 (Print)).

40. Youd E, Moore LA-OX. COVID-19 autopsy in people who died in community settings: the first series. LID - jclinpath-2020-206710 [pii] LID - 10.1136/jclinpath-2020-206710 [doi]. (1472-4146 (Electronic)).

41. Suess C, Hausmann R. Gross and histopathological pulmonary findings in a COVID-19 associated death during self-isolation. (1437-1596 (Electronic)).

42. Bruni A, Garofalo E, Zuccalà V, Currò G, Torti C, Navarra G, et al. Histopathological findings in a COVID-19 patient affected by ischemic gangrenous cholecystitis. (1749-7922 (Electronic)).

43. Colmenero IA-OX, Santonja C, Alonso-Riaño M, Noguera-Morel LA-O, Hernández-MartÎn AA-O, Andina D, et al. SARS-CoV-2 endothelial infection causes COVID-19 chilblains: histopathological, immunohistochemical and ultrastructural study of seven paediatric cases. LID - 10.1111/bjd.19327 [doi]. (1365-2133 (Electronic)).

44. Beigmohammadi MT, Jahanbin B, Safaei M, Amoozadeh L, Khoshavi M, Mehrtash V, et al. Pathological Findings of Postmortem Biopsies From Lung, Heart, and Liver of 7 Deceased COVID-19 Patients. (1940-2465 (Electronic)).

45. Heinrich FA-O, Sperhake JP, Heinemann A, Mushumba H, Lennartz M, Nörz D, et al. Germany’s first COVID-19 deceased: a 59-year-old man presenting with diffuse alveolar damage due to SARS-CoV-2 infection. (1432-2307 (Electronic)).

46. Wang C, Xie J, Zhao L, Fei X, Zhang H, Tan Y, et al. Alveolar macrophage dysfunction and cytokine storm in the pathogenesis of two severe COVID-19 patients. EBioMedicine. 2020;57.

47. Reichard RA-O, Kashani KB, Boire NA, Constantopoulos E, Guo Y, Lucchinetti CF. Neuropathology of COVID-19: a spectrum of vascular and acute disseminated encephalomyelitis (ADEM)-like pathology. (1432-0533 (Electronic)).

48. Yao XH, Li TY, He ZC, Ping YF, Liu HW, Yu SC, et al. [A pathological report of three COVID-19 cases by minimal invasive autopsies]. (0529-5807 (Print)).

49. Menter T, Haslbauer JD, Nienhold R, Savic S, Hopfer H, Deigendesch N, et al. Postmortem examination of COVID-19 patients reveals diffuse alveolar damage with severe capillary congestion and variegated findings in lungs and other organs suggesting vascular dysfunction. Histopathology. 2020;77(2):198–209.

50. Edler C, Schröder AS, Aepfelbacher M, Fitzek A, Heinemann A, Heinrich F, et al. Dying with SARS-CoV-2 infection-an autopsy study of the first consecutive 80 cases in Hamburg, Germany. (1437-1596 (Electronic)).

51. Remmelink M, De Mendoca R, Haene N, De Clercq S, Verocq C, Lebrun L, et al. Unspecific postmortem findings despite multiorgan 1 viral spread in COVID-19 patients. medRxiv. 2020:2020.05.27.20114363.

52. Lax SF, Skok K, Zechner P, Kessler HH, Kaufmann N, Koelblinger C, et al. Pulmonary Arterial Thrombosis in COVID-19 With Fatal Outcome : Results From a Prospective, Single-Center, Clinicopathologic Case Series. (1539-3704 (Electronic)).

53. Magro C, Mulvey JJ, Berlin D, Nuovo G, Salvatore S, Harp J, et al. Complement associated microvascular injury and thrombosis in the pathogenesis of severe COVID-19 infection: A report of five cases. (1878-1810 (Electronic)).

54. Bryce C, Grimes Z, Pujadas E, Ahuja S, Beasley MB, Albrecht R, et al. Pathophysiology of SARS-CoV-2: targeting of endothelial cells renders a complex disease with thrombotic microangiopathy and aberrant immune response. The Mount Sinai COVID-19 autopsy experience. medRxiv. 2020:2020.05.18.20099960.

55. Fitzek A, Sperhake J, Edler C, Schröder AS, Heinemann A, Heinrich F, et al. Evidence for systematic autopsies in COVID-19 positive deceased. Rechtsmedizin. 2020;30(3):184–9.

56. Adachi T Fau - Chong J-M, Chong Jm Fau - Nakajima N, Nakajima N Fau - Sano M, Sano M Fau - Yamazaki J, Yamazaki J Fau - Miyamoto I, Miyamoto I Fau - Nishioka H, et al. Clinicopathologic and Immunohistochemical Findings from Autopsy of Patient with COVID-19, Japan. LID - 10.3201/eid2609.201353 [doi]. (1080-6059 (Electronic)).

57. Flikweert AW, Grootenboers M, Yick DCY, du Mée AWF, van der Meer NJM, Rettig TCD, et al. Late histopathologic characteristics of critically ill COVID-19 patients: Different phenotypes without evidence of invasive aspergillosis, a case series. (1557-8615 (Electronic)).

58. Ackermann M, Verleden SE, Kuehnel M, Haverich A, Welte T, Laenger F, et al. Pulmonary Vascular Endothelialitis, Thrombosis, and Angiogenesis in Covid-19. New England Journal of Medicine. 2020;383(2):120–8.

59. Schaefer IA-O, Padera RF, Solomon IA-O, Kanjilal S, Hammer MM, Hornick JL, et al. In situ detection of SARS-CoV-2 in lungs and airways of patients with COVID-19. (1530-0285 (Electronic)).

60. Bösmüller HA-OX, Traxler S, Bitzer M, Häberle H, Raiser W, Nann D, et al. The evolution of pulmonary pathology in fatal COVID-19 disease: an autopsy study with clinical correlation. (1432-2307 (Electronic)).

61. Sonzogni AA-O, Previtali GA-O, Seghezzi MA-O, Grazia Alessio MA-O, Gianatti AA-O, Licini L, et al. Liver histopathology in severe COVID 19 respiratory failure is suggestive of vascular alterations. LID - 10.1111/liv.14601 [doi]. (1478-3231 (Electronic)).

62. Wang Y, Liu S, Liu H, Li W, Lin F, Jiang L, et al. SARS-CoV-2 infection of the liver directly contributes to hepatic impairment in patients with COVID-19. LID - S0168-8278(20)30294-4 [pii] LID - 10.1016/j.jhep.2020.05.002 [doi]. (1600-0641 (Electronic)).

63. Ducloyer MA-O, Gaborit B, Toquet C, Castain L, Bal A, Arrigoni PP, et al. Complete postmortem data in a fatal case of COVID-19: clinical, radiological and pathological correlations. (1437-1596 (Electronic)).

64. Kantonen J, Mahzabin S, Mäyränpää MI, Tynninen O, Paetau A, Andersson N, et al. Neuropathologic features of four autopsied COVID-19 patients. LID - 10.1111/bpa.12889 [doi]. (1750-3639 (Electronic)).

65. CÎrstea AE, Buzulicâ Rl Fau - Pirici D, Pirici D Fau - Ceauşu MC, Ceauşu Mc Fau - Iman RV, Iman Rv Fau - Gheorghe OM, Gheorghe Om Fau - Neamţu SD, et al. Histopathological findings in the advanced natural evolution of the SARS-CoV-2 infection. (1220-0522 (Print)).

66. Schwensen HA-OX, Borreschmidt LK, Storgaard M, Redsted S, Christensen S, Madsen LB. Fatal pulmonary fibrosis: a post-COVID-19 autopsy case. LID - jclinpath-2020- 206879 [pii] LID - 10.1136/jclinpath-2020-206879 [doi]. (1472-4146 (Electronic)).

67. Santoriello D, Khairallah P, Bomback AS, Xu K, Kudose SA-O, Batal I, et al. Postmortem Kidney Pathology Findings in Patients with COVID-19. (1533-3450 (Electronic)).

68. Salerno M, Sessa F, Piscopo A, Montana A, Torrisi M, Patanè F, et al. No Autopsies on COVID-19 Deaths: A Missed Opportunity and the Lockdown of Science. LID - 10.3390/jcm9051472 [doi] LID - 1472. (2077-0383 (Print)).

69. Barranco R, Ventura F. The role of forensic pathologists in coronavirus disease 2019 infection: The importance of an interdisciplinary research. Medicine, Science and the Law. 2020;60(3):237–8.

70. Fineschi V, Aprile A, Aquila I, Arcangeli M, Asmundo A, Bacci M, et al. Management of the corpse with suspect, probable or confirmed COVID-19 respiratory infection - Italian interim recommendations for personnel potentially exposed to material from corpses, including body fluids, in morgue structures and during autopsy practice. (1591- 951X (Electronic)).

71. Dijkhuizen LGM, Gelderman HT, Duijst W. Review: The safe handling of a corpse (suspected) with COVID-19. (1878-7487 (Electronic)).

72. Ng DL, Al Hosani F, Keating MK, Gerber SI, Jones TL, Metcalfe MG, et al. Clinicopathologic, Immunohistochemical, and Ultrastructural Findings of a Fatal Case of Middle East Respiratory Syndrome Coronavirus Infection in the United Arab Emirates, April 2014. Am J Pathol. 2016;186(3):652–8.

73. Nicholls JM, Poon Ll Fau - Lee KC, Lee Kc Fau - Ng WF, Ng Wf Fau - Lai ST, Lai St Fau - Leung CY, Leung Cy Fau - Chu CM, et al. Lung pathology of fatal severe acute respiratory syndrome. (0140-6736 (Print)).

74. Harms PW, Schmidt LA, Smith LB, Newton DW, Pletneva MA, Walters LL, et al. Autopsy Findings in Eight Patients With Fatal H1N1 Influenza. American Journal of Clinical Pathology. 2010;134(1):27–35.

75. Balraam KA-O, Sidhu AA-OX, Srinivas VA-O. Interesting postmortem findings in a H1N1 influenza-positive pneumonia patient. (2236-1960 (Print)).

76. Nakajima N, Sato Y Fau - Katano H, Katano H Fau - Hasegawa H, Hasegawa H Fau - Kumasaka T, Kumasaka T Fau - Hata S, Hata S Fau - Tanaka S, et al. Histopathological and immunohistochemical findings of 20 autopsy cases with 2009 H1N1 virus infection. (1530- 0285 (Electronic)).

77. Skálová H Fau - Povýšil C, Povýšil C Fau - Hofmanová J, Hofmanová J Fau - Goldová B, Goldová B Fau - Jakša R, Jakša R Fau - Jandová K, Jandová K Fau - Galko J, et al. Histopathological autoptic findings in 8 patients with pandemic influenza A (H1N1) pneumonia. (1210-7875 (Print)).

78. Sawalha K, Abozenah M, Kadado AJ, Battisha A, Al-Akchar M, Salerno C, et al. Systematic review of COVID-19 related myocarditis: Insights on management and outcome. LID - S1553-8389(20)30497-8 [pii] LID - 10.1016/j.carrev.2020.08.028 [doi]. (1878-0938 (Electronic)).

79. Lang ZW, Zhang Lj Fau - Zhang S-J, Zhang Sj Fau - Meng X, Meng X Fau - Li J-Q, Li Jq Fau - Song C-Z, Song Cz Fau - Sun L, et al. A clinicopathological study of three cases of severe acute respiratory syndrome (SARS). (0031-3025 (Print)).

80. Chong Py Fau - Chui P, Chui P Fau - Ling AE, Ling Ae Fau - Franks TJ, Franks Tj Fau - Tai DYH, Tai Dy Fau - Leo YS, Leo Ys Fau - Kaw GJL, et al. Analysis of deaths during the severe acute respiratory syndrome (SARS) epidemic in Singapore: challenges in determining a SARS diagnosis. (1543-2165 (Electronic)).

81. Gu J, Gong E Fau - Zhang B, Zhang B Fau - Zheng J, Zheng J Fau - Gao Z, Gao Z Fau - Zhong Y, Zhong Y Fau - Zou W, et al. Multiple organ infection and the pathogenesis of SARS. (0022-1007 (Print)).

82. Wu VC, Hsueh Pr Fau - Lin W-C, Lin Wc Fau - Huang J-W, Huang Jw Fau - Tsai H-B, Tsai Hb Fau - Chen Y-M, Chen Ym Fau - Wu K-D, et al. Acute renal failure in SARS patients: more than rhabdomyolysis. (0931-0509 (Print)).

83. Chau T-N, Lee K-C, Yao H, Tsang T-Y, Chow T-C, Yeung Y-C, et al. SARS-associated viral hepatitis caused by a novel coronavirus: Report of three cases. Hepatology. 2004;39(2):302–10.

84. Wong RS, Wu A Fau - To KF, To Kf Fau - Lee N, Lee N Fau - Lam CWK, Lam Cw Fau - Wong CK, Wong Ck Fau - Chan PKS, et al. Haematological manifestations in patients with severe acute respiratory syndrome: retrospective analysis. (1756-1833 (Electronic)).

85. Neumann B, Schmidbauer ML, Dimitriadis K, Otto S, Knier B, Niesen WD, et al. Cerebrospinal fluid findings in COVID-19 patients with neurological symptoms. (1878-5883 (Electronic)).

86. Xu J, Zhong S Fau - Liu J, Liu J Fau - Li L, Li L Fau - Li Y, Li Y Fau - Wu X, Wu X Fau - Li Z, et al. Detection of severe acute respiratory syndrome coronavirus in the brain: potential role of the chemokine mig in pathogenesis. (1537-6591 (Electronic)).

